# Utilizing Machine Learning Models to Predict Acute Kidney Injury in Septic Patients from MIMIC-III Database

**DOI:** 10.1101/2025.03.11.25323787

**Authors:** Aleyeh Roknaldin, Junyi Fan, Shuheng Chen, Elham Pishgar, Greg Placencia, Kamiar Alaei, Maryam Pishgar

**Affiliations:** Department of Industrial and Systems Engineering, University of Southern California, Los Angeles, CA, United States of America; Colorectal Research Center, Iran University of Medical Sciences, Tehran, Iran; Department of Industrial and Manufacturing Engineering, California State Polytechnic University, Pomona, Los Angeles, CA, United States of America; Department of Health Science, California State University, Long Beach, Long Beach, CA, United States of America

## Abstract

**Background:** Sepsis is a severe condition that causes the body to respond incorrectly to an infection. This reaction can subsequently cause organ failure, a major one being acute kidney injury (AKI). For septic patients, approximately 50% develop AKI, with a mortality rate above 40%. Creating models that can accurately predict AKI based on specific qualities of septic patients is crucial for early detection and intervention.

**Methods:** Using medical data from septic patients during intensive care unit (ICU) admission from the Medical Information Mart for Intensive Care 3 (MIMIC-III) dataset, we extracted 3301 patients with sepsis, with 73% of patients developing AKI. The data was randomly divided into a training set (n = 1980, 40%), a test set (n = 661, 10%), and a validation set (n = 660, 50%). The proposed model was logistic regression, and it was compared against five baseline models: XGBoost, K Nearest Neighbors (KNN), Support Vector Machines (SVM), Random Forest (RF) and LightGBM. Area Under the Curve (AUC), Accuracy, F1-Score, and Recall were calculated for each model.

**Results:** After analysis, we were able to select 23 features to include in our model, the top features being urine output, maximum bilirubin, minimum bilirubin, weight, maximum blood urea nitrogen, and minimum estimated glomerular filtration rate. The logistic regression model performed the best, achieving an AUC score of 0.887 (95% CI: [0.861-0.915]), an accuracy of 0.817, an F1 score of 0.866, a recall score of 0.827, and a Brier score of 0.13.

**Conclusion:** Compared to the best existing literature in this field, our model achieved an 8.57% improvement in AUC while using 13 less variables, showcasing its effectiveness in determining AKI in septic patients. While the features selected for predicting AKI in septic patients are similar to previous literature, the top features that influenced our model’s performance differ.

## Introduction

Acute kidney injury (AKI) is an intricate medical condition in which the kidneys are ineffective at filtering waste from the blood. This decline in renal function leads to the accumulation of waste, causing the kidneys to be unable to maintain the balance of electrolyte, acid-base, and water-all of which can lead to other organ problems such as cardiovascular, gastrointestinal, and neurological complications [1, 2]. Despite medical advances in the treatment of AKI, recent studies have found that the mortality rate related to AKI is about 23% [3].

The most common cause of AKI is sepsis [4]. Sepsis-associated AKI is among the most fatal, with 53% of septic patients developing AKI, and accounts for a mortality rate of up to 44% [5]. Sepsis-associated AKI can also lead to longer hospital stays, increased development of long-term disabilities, and lower quality of life [6]. Early detection in high-risk septic patients can lead to the reversal of renal failure in the early stages, while also improving patient care in the Intensive Care Unit (ICU) and recovery through medical interventions such as Renal Replacement Therapy [7]. It is imperative to understand the predictors associated with AKI in septic patients, but the complexity of the pathophysiology in sepsis-associated AKI makes it difficult to timely intervention and prevent renal injury. In general, those who seek medical attention have already developed AKI. Currently, a prominent measure for the diagnosis of AKI is based on increased creatinine levels and a decrease in urine production [8] [4]. However, studies have shown that these biomarkers may be nonspecific and require exploring other indicators [7]. When assessing risk factors, [9] found that septic shock, hypertension, the use of vasopressors, and mechanical ventilation were among the main determinants that could increase the chance of AKI associated with sepsis.

To mitigate the challenges in early detection of AKI in septic patients, several studies have investigated various machine learning and statistical methods to construct predictive models [10, 11]. The models utilize specific features or indicators to predict which septic patients develop AKI. In addition to machine learning models, other studies have also used scoring systems such as the simplified acute physiology score (SAPS) II and the sequential organ failure assessment (SOFA), but resulted in poor performance primarily due to low specificity and sensitivity [5].

Our study contributes to the prediction of AKI risk employing a streamlined set of only 23 characteristics, separating our model from the extensive set of variables commonly used in models found in the literature. Our features were carefully selected to enhance the interpretability of the model and reduce overfitting. This focused feature selection aligns with the best practices in sepsis mortality studies, which prioritize both predictive precision and clinical relevance. By adopting a methodology that mirrors rigorous research on sepsis mortality, our approach aims to balance precision with practical usability in clinical settings. Furthermore, the predictive model was developed in accordance with the Transparent Reporting of Individual Prognostic or Diagnostic Multivariate Predictive Model (TRIPOD) guidelines, ensuring transparency and robust cross-cohort comparability.

The structure of our study, from data extraction to validation, mirrors that of recent high-impact research on sepsis mortality, ensuring robust cross-cohort comparability. Additionally, we use Logistic Regression as the primary model due to its balance of performance and interpretability, demonstrating strong results in predictive metrics, including AUC and Brier scores.

In summary, this study advances the prediction of AKI risk in septic patients by adopting a minimal yet effective set of characteristics of 23 variables and reducing the complexity of the model while maintaining high predictive power. By prioritizing interpretability and alignment with established clinical guidelines (TRIPOD), our approach not only improves transparency, but also enhances the model’s clinical utility. Logistic regression was chosen for its balanced performance and ease of interpretation, demonstrating strong predictive metrics such as AUC, accuracy, and Brier scores, which support its application in real-world clinical decision-making.

## Methods

### Data Source

The MIMIC-III database, sourced from the ICU at Beth Israel Deaconess Medical Center in Boston, contains extensive health data on over 40,000 patients from 2001 to 2012 [12]. This robust data set features detailed records from more than 58,000 hospitalizations, including physiological signals, medications, lab results, diagnostic data, and medical summaries. Focuses on a diverse range of patients in the ICU, providing vital information for clinical decision-making and patient outcomes. The database was crucial for our study as it provided real patient data used to develop our models. The MIMIC-III dataset used in this study is publicly available at https://physionet.org/content/mimiciii/1.4/.

### Participant Selection

Our focus was on patients who currently have sepsis. The flow chart in Figure 1 gives a breakdown of the screening and classification requirements that are carried out to reach the final data set. Initially, we filtered the MIMIC-III database using the unique ICD-9 codes 99591, 99592, and 78552 to find patients with sepsis. This left us with 6138 septic patients. We then excluded patients not within the age range of 18 to 89 years, patients with multiple admissions, patients who stayed in the ICU for less than 48 hours, and patients who had more than 20% missing values. Lastly, patients with incomplete information were also eliminated. Initially, we encountered 0.7% of our final dataset with more than 11 missing values. Given that this was less than 1% of the data, we removed these rows, assuming that it would not affect our result. We finalized the data set for 3301 septic patients, 2410 of whom developed AKI and 891 of whom did not.

**Figure 1.**
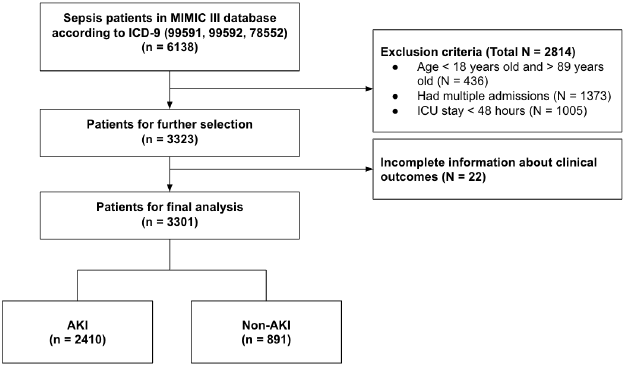
Flow chart of patient selection from MIMIC-III dataset.

### Data Extraction

Patient data from the first 24 hours after admission were extracted from the MIMIC-III database to capture the early clinical status. Demographic information provided context on patient variability and potential risk factors, while comorbidities highlighted underlying conditions that could influence disease progression. Vital signs reflected physiological stability, offering key indicators of cardiovascular, respiratory, and metabolic function. Laboratory values provided biochemical information on organ performance, electrolyte balance, and coagulation status. In addition, therapeutic interventions such as mechanical ventilation and vasopressor use served as proxy for the severity of the disease. For continuously measured variables, maximum and minimum values were recorded to account for physiological fluctuations. Detailed indicators are shown in the following table.

To reduce bias caused by missing data, features with more than 20 missing values % were excluded from the final cohort. The missing values in other features were imputed using the multiple imputation (MI) method. Multiple Imputation (MI) is a statistical method widely used for handling missing data. Address missing values by generating multiple imputed datasets, each with the missing values being replaced with plausible values. These data sets are then synthetically analyzed, obtaining more accurate estimates of the missing values.

### Feature Selection

After an initial filtering of septic patients and grouping them into AKI and non-AKI, we started with 50 characteristics. To determine the main characteristics, we used a correlation-based method. Each characteristic *X*_*i*_ was evaluated for its correlation with the target variable *Y* (AKI presence: 1 for AKI, 0 for non-AKI) using the Pearson correlation coefficient:

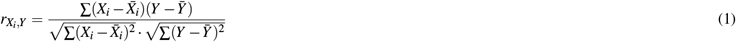

We selected features with an absolute correlation coefficient within the range:

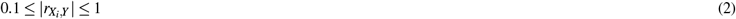

leaving us with the 23 most correlated features to AKI. All features satisfying 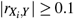 were considered for further analysis.Table 1 shows the 23 features selected, which are (i) age: patient age when enter ICU; (ii) minimum heart rate: patient’s minimum recorded heart rate; (iii) minimum temperature: patient’s minimum recorded temperature; (iv) minimum oxygen saturation (SpO2): patient’s minimum recorded oxygen saturation; (v) minimum systolic blood pressure (SysBP): patient’s minimum recorded systolic blood pressure; (vi) minimum diastolic blood pressure (DiasBP): patient’s minimum recorded diastolic blood pressure; (vii) minimum bilirubin: patient’s minimum recorded bilirubin from lab results; (viii) maximum bilirubin: patient’s maximum recorded bilirubin from lab results; (ix) minimum aniongap: patient’s minimum recorded aniongap from lab results; (x) maximum aniongap: patient’s maximum recorded aniongap from lab results; (xi) minimum potassium: patient’s minimum recorded potassium level from lab results; (xii) maximum potassium: patient’s maximum recorded potassium level from lab results; (xiii) minimum lactate: patient’s minimum recorded lactate level from lab results; (xiv) maximum lactate: patient’s maximum recorded lactate level from lab results; (xv) minimum creatinine: patient’s minimum recorded creatinine level from lab results; (xvi) maximum creatinine: patient’s maximum recorded creatinine level from lab results; (xvii) minimum blood urea nitrogen (BUN): patient’s minimum recorded blood urea nitrogen from lab results;(Xviii) maximum blood urea nitrogen (BUN): patient’s maximum recorded blood urea nitrogen from lab results; (xix) urine output: patient’s recorded urine output; (xx) vasopressor (vaso): whether patient took vasopressor or not; (xxi) weight: patient’s weight at ICU admission; (xxii) minimum estimated glomerular filtration rate (eGFR): patient’s minimum recorded estimated glomerular filtration rate from lab results; and (xxiii) mechanical ventilation: whether a patient uses mechanical ventilation or not.

**Table 1.** Feature Category Table.

| Category | Feature | Unit |
| --- | --- | --- |
| Demographic | Age<br>Weight | years<br>lbs |
| Vital Signs | Minimum Heart Rate<br>Minimum Temperature<br>Minimum Oxygen Saturation (SpO2)<br>Minimum Systolic Blood Pressure (SysBP)<br>Minimum Diastolic Blood Pressure (DiasBP)<br>Urine Output | per minute<br>°C<br>%<br>mmHg<br>mmHg<br>ml |
| Laboratory Results | Minimum Bilirubin<br>Maximum Bilirubin<br>Minimum Anion Gap<br>Maximum Anion Gap<br>Minimum Potassium<br>Maximum Potassium<br>Minimum Lactate<br>Maximum Lactate<br>Minimum Creatinine<br>Maximum Creatinine<br>Minimum Blood Urea Nitrogen (BUN)<br>Maximum Blood Urea Nitrogen (BUN)<br>Minimum Estimated Glomerular Filtration Rate (eGFR) | mg/dL<br>mg/dL<br>mmol/L<br>mmol/L<br>mEq/L<br>mEq/L<br>mmol/L<br>mmol/L<br>mg/dL<br>mg/dL<br>mg/dL<br>mg/dL<br>- |
| Interventions | Mechanical Ventilation<br>Vasopressor (Vaso) | Yes/No<br>Yes/No |

Our main characteristics were urine output, minimum eGFR, maximum eGFR, minimum SysBP, mechanical ventilation, vasopressor, weight, and minimum creatinine. The existing literature includes urine output, mechanical ventilation, vasopressor, and creatinine as the main characteristics, but the high correlation of minimum eGFR, maximum eGFR, minimum SysBP, and weight to predict AKI in septic patients is unique to our model.

Urine output is one of the most common signs of AKI. Studies have been done to show the ideal amount and time to collect urine sample measurements [13]. A decrease in urine content is typically a strong sign of AKI, making its importance as a characteristic a high factor in prediction. Regarding eGFR levels, it has been seen to decrease after patients are diagnosed with AKI [14]. Although there is still ongoing research on the effect of eGFR and AKI, it is a notable feature that should be considered for prediction. Systolic blood pressure is another indicator that should be considered when determining an AKI diagnosis because, based on previous studies, it is vital for organ perfusion, where blood flow provides oxygen and nutrients to the organs [15, 16]. Vasopressor is a drug that is used to increase blood pressure. It can be used to regulate hypotension by constricting blood vessels [17]. In our dataset, it was recorded whether a patient took vasopressor or not. Weight is another significant factor in the prediction of AKI. Studies have stated that obesity is associated with an increased risk of AKI in areas such as acute respiratory distress syndrome or after cardiac surgery [18, 19]. In a recent study involving critically ill septic patients in the ICU, obesity patients were found to have a higher chance of developing early sepsis-associated AKI [20]. The literature supports our findings on the characteristics that will be used in the development of a model to predict AKI in septic patients.

### Statistical Analysis for AKI and Non-AKI Patients

To compare the characteristics of patients with AKI and non-AKI, we used statistical tests with an alpha set to 0.05 to compare the two cohorts and see if there are statistically significant differences. The results can be found in Table **??**. Quantitative values were compared using a T test and qualitative values were compared using a chi-squared test. All features were statistically significant between AKI patients and non-AKI patients.

### Model Development

Our filtered data set was imbalanced between AKI patients and non-AKI patients. There were 2 410 AKI patients and 891 non-AKI patients. To fix this, we used the synthetic minority over-sampling technique (SMOTE), which creates additional data from the minority class using the K Nearest Neighbors approach [21]. SMOTE was added to all the features that have been selected to balance the data and prevent overfitting. The data was then divided into a training, testing, and validation cohort. The six models used in this study were Logistic Regression (LR), XGBoost, K Nearest Neighbors (KNN), Support Vector Machines (SVM), Random Forest (RF), and LightGBM.

Logistic regression has been studied to have comparable performance results when compared to machine learning models and has been used in studies for predicting AKI [22, 23]. For its parameters, we used a C of 100, a maximum iteration of 200, and a penalty of ‘l2’. XGBoost has been used in previous studies for predicting AKI in patients with specific prior health conditions [24, 25]. For the parameters, we used the ‘binary:logistic’ objective, a ‘reg_lambda’ of 100, a ‘reg_alpha’ of 120, and a ‘max_depth’ of 2. KNN is another machine learning algorithm that utilizes classification and has been used for medical predictions, such as predicting development of AKI for heart failure patients [26, 27]. For its parameters, we used an n_neighbors of 40. SVM is another well-known machine learning model known to provide high accuracy [28]. For SVM, we used a C of 0.1 and a gamma of 0.02. Random forest is a widely utilized classification machine learning model that has been used in several acute kidney injury prediction studies [29, 30]. We used an n_estimator of 150, a max_depth of 12, a min_samples_split of 128, and a min_samples_leaf of 10. LightGBM is another commonly used machine learning model in the literature for predicting AKI in septic patients. We used the following hyperparameters: *λ*_*l*1_ = 1, *λ*_*l*2_ = 1, num_leaves = 20, max_depth = 4, random_state = 42, learning_rate = 0.01, n_estimators = 1050, and verbose =− 1.

The best model was determined on the basis of its respective AUC value. Although we also looked at the accuracy, F1 score, and recall of each model, we decided to use AUC as the metric of comparison since it is more suitable for handling unbalanced datasets and when comparing multiple models.

For this study, the data was imported to BigQuery and extracted using SQL. Data preprocessing, feature selection, and training were performed using Python 3.10.12.

### Shapley Analysis

To interpret the predictive power of each characteristic within our model, we employed Shapley analysis. Shapley values assign a contribution score to each feature based on its marginal impact on the model’s output, taking into account all possible feature combinations. This approach provides a comprehensive measure of the importance of features that is both rigorous and interpretable, making it especially suited to healthcare applications where understanding the influence of each variable is crucial. Shapley analysis helps us identify the most critical predictors, such as urine output, bilirubin, and blood urea nitrogen levels. These values are particularly useful for determining how each characteristic individually, as well as in combination with others, influences the likelihood of AKI, thus allowing for more informed clinical insights.

The analysis was chosen because of its ability to illustrate the contributions of the characteristics in a transparent way, which is consistent with the need for interpretability in clinical decision making tools [31]. This analysis enabled us to pinpoint the most impactful characteristics, visualized in our results section, that support the application of our model in real world settings where clinicians can focus on key indicators of risk of AKI.

## Results

### Training and Validation Comparison

After an initial filtering of patients who did not meet the analysis requirements, we were left with 3301 patients and 56 characteristics. After narrowing our characteristics down to 23 using correlation, the data was then randomly divided into 40% for the training set, 10% for the test set, and 50% for the validation set, resulting in 1980 patients for training, 661 patients for testing, and 660 for validation. Table 2 gives a comprehensive comparison between the training and validation sets. The comparison of the training and validation set is to confirm that our null hypothesis of no difference between feature distributions holds.

**Table 2.** Averages and P-values between features of the training and validation set.

| Category | Training Set (N = 1980) | Validation Set (N = 660) | P-value |
| --- | --- | --- | --- |
| **Demographic** |  |  |  |
| Age (years) | 64.8 | 64.1 | 0.303 |
| Weight (lbs) | 83.4 | 84.0 | 0.647 |
| **Vital Signs** |  |  |  |
| Minimum Heart Rate (per minute) | 62.7 | 62.1 | 0.358 |
| Minimum Temperature (Celsius) | 35.5 | 35.5 | 0.548 |
| Minimum Oxygen Saturation (SpO2) (%) | 80.0 | 80.0 | 0.946 |
| Minimum Systolic Blood Pressure (SysBP) (mmHg) | 71.1 | 72.2 | 0.198 |
| Minimum Diastolic Blood Pressure (DiasBP) (mmHg) | 32.6 | 33.1 | 0.290 |
| Urine Output (ml) | 1576 | 1696 | 0.085 |
| **Laboratory Results** |  |  |  |
| Minimum Bilirubin (mg/dL) | 2.62 | 2.51 | 0.627 |
| Maximum Bilirubin (mg/dL) | 3.05 | 3.00 | 0.823 |
| Minimum Anion Gap (mmol/L) | 13.6 | 13.6 | 0.936 |
| Maximum Anion Gap (mmol/L) | 17.7 | 17.7 | 0.796 |
| Minimum Potassium (mEq/L) | 3.73 | 3.73 | 0.920 |
| Maximum Potassium (mEq/L) | 4.66 | 4.66 | 0.907 |
| Minimum Lactate (mmol/L) | 1.84 | 1.86 | 0.717 |
| Maximum Lactate (mmol/L) | 3.36 | 3.32 | 0.767 |
| Minimum Creatinine (mg/dL) | 1.73 | 1.72 | 0.885 |
| Maximum Creatinine (mg/dL) | 2.17 | 2.15 | 0.764 |
| Minimum Blood Urea Nitrogen (BUN) (mg/dL) | 33.7 | 32.9 | 0.453 |
| Maximum Blood Urea Nitrogen (BUN) (mg/dL) | 40.6 | 39.7 | 0.488 |
| Minimum Estimated Glomerular Filtration Rate (eGFR) | 68.0 | 69.0 | 0.520 |
| **Interventions** |  |  |  |
| Mechanical Ventilation (count) | 1425 | 465 | 0.485 |
| Vasopressor (Vaso) (count) | 1104 | 367 | 0.981 |

### Model Performance Evaluation

After running each of the six models, we have documented our results in Tables 3 and 4. The Logistic Regression model had a training set AUC of 0.898 (95% CI = [0.888 - 0.909]), a validation set AUC of 0.857 (95% CI = [0.829 – 0.885]), and a testing set AUC of 0.887 (95% CI = [0.861 - 0.915]). Regarding the other five models, LightGBM 0.885, SVM had an AUC of 0.876, random forest 0.867, XGBoost 0.862, and KNN 0.725.

**Table 3.** Logistic Regression Model training, testing, and validation sets evaluation metrics.

| Models | AUC | AUC 95% CI | Accuracy |
| --- | --- | --- | --- |
| Proposed model performance on test set | 0.887 | [0.861-0.915] | 0.817 |
| Proposed model performance on validation set | 0.857 | [0.829–0.885] | 0.791 |
| Proposed model performance on training set | 0.898 | [0.888-0.909] | 0.819 |

**Table 4.** Model Evaluation Metrics.

| Models | AUC | AUC 95% CI | Accuracy | F1 Score | Recall |
| --- | --- | --- | --- | --- | --- |
| Logistic Regression | 0.887 | [0.861-0.915] | 0.817 | 0.867 | 0.827 |
| LightGBM | 0.885 | [0.860-0.911] | 0.815 | 0.869 | 0.853 |
| SVM | 0.876 | [0.844-0.904] | 0.797 | 0.849 | 0.794 |
| Random Forest | 0.867 | [0.839-0.895] | 0.790 | 0.848 | 0.819 |
| XGBoost | 0.862 | [0.834-0.891] | 0.796 | 0.777 | 0.739 |
| KNN | 0.725 | [0.683-0.764] | 0.696 | 0.777 | 0.739 |

Figure 2 shows the AUC curves of each of the 6 models tested. Logistic regression exhibits the closest shape to an ideal AUC curve, showcasing its superior performance amongst the other models.

**Figure 2.**
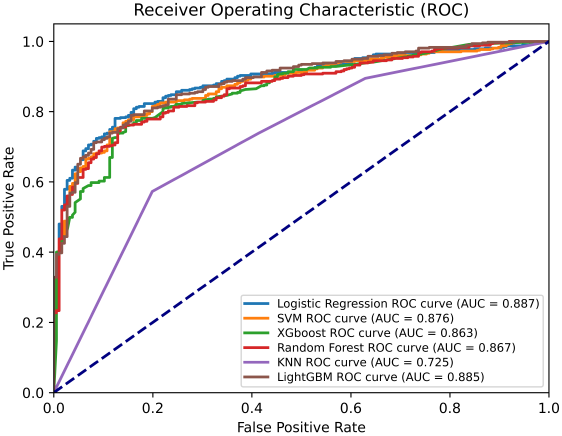
ROC curves of Baseline and Proposed models.

In addition, we applied rigorous calibration techniques to ensure the reliability of the predicted probabilities of each model. Model calibration was assessed using the Brier score, which quantifies the mean squared difference between predicted probabilities and actual outcomes, a lower score indicating better calibration. Among the models tested, logistic regression achieved the lowest Brier score (0.134), followed by LightGBM (0.144), SVM (0.141), Random Forest (0.143), XGBoost (0.147) and KNN (0.229), demonstrating its superior calibration performance. To further refine the models, we applied isotonic regression, a non-parametric technique, to adjust the probability estimates. As shown in Figure 3, the predicted probabilities align closely with the diagonal line, indicating an effective calibration. The combination of a low Brier score and a high AUC underscores the accuracy and reliability of logistic regression.

**Figure 3.**
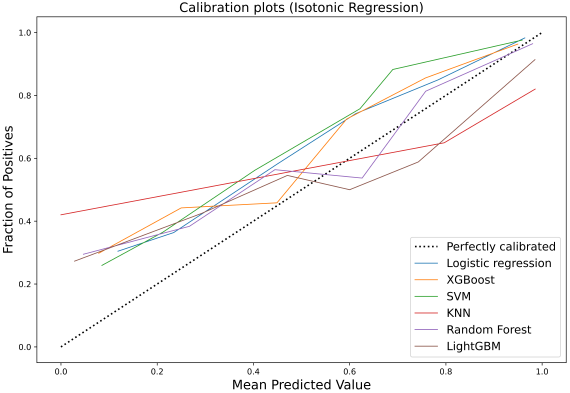
Calibration curves of the 6 models used: Logistic Regression, SVM, XGBoost, Random Forest, KNN, and LightGBM.

### Shapley Analysis for Logistic Regression

Shapley analysis is a common measure to see which features contribute the most to the model’s overall performance. Since Logistic Regression was our best model, we applied Shapley analysis to see which features most affected the outcome of our testing set.

Figure 4 visualizes the Shapley analysis for our Logistic Regression model. Urine output, maximum bilirubin, minimum bilirubin, and weight are the most important features used in the Logistic Regression prediction model. From Figure 5, we see that urine output has the most significant impact on the Logistic Regression model. Low urine output increases the risk of AKI, displayed by its negative SHAP value. This is consistent with clinical understanding where reduced urine output is a key indicator of acute kidney injury (AKI) [32]. Elevated bilirubin levels push predictions toward higher risk, reflecting liver dysfunction or sepsis-induced cholestasis, conditions often seen in critical care patients [33].

**Figure 4.**
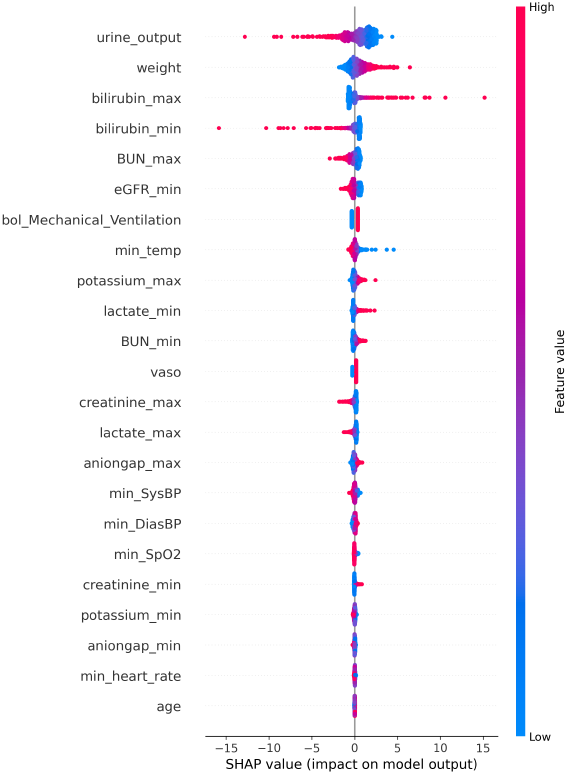
Shapley Analysis of Logistic Model features.

**Figure 5.**
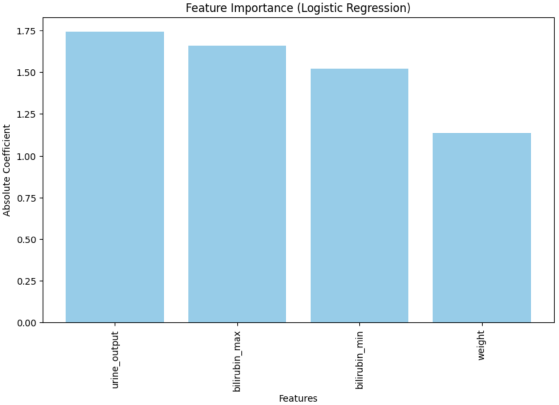
Feature importance for logistic regression test set.

Compared to the importance of the original feature, it is interesting to note that bilirubin now plays an important role in the predictive process. While in the original feature correlation results it was not among the top features, it now holds a higher predictive power in the Logistic Regression model. This highlights the importance of performing additional analysis to understand what specific features the model looks at to make predictions.

## Discussion

Our model aims to predict acute kidney injury in septic patients. Among all the models we have trained, the proposed Logistic Regression model shows an impressive outcome with an AUC of 0.887, which is only 0.23% higher than the second-best model trained, LightGBM. These results exceed the general pattern seen in previous literature.

In the field of predicting acute kidney injury, several studies have generated proper outcomes using different methods, including XGBoost, LightGBM, and Nomogram [5], [34–37]. Compared to the best literature model by [5], our model shows a significant improvement. The best literature model uses XGBoost, while ours uses Logistic Regression. Our model shows an 8.56% improvement in AUC. Additionally, our AUC values contain precise confidence intervals, ensuring reliability of our results. While the best existing model used 36 variables, our model was able to use 13 less variables and achieve a higher AUC score. The lower feature count prevents issues such as overfitting and highlights the efficiency of our model. This is crucial when working with sparse medical data to ensure accurate predictions. Like the best existing literature, we also utilized a training, validation, and test set. Regarding the features used, we have three features not included in the model of the current best literature: weight, minimum heart rate, and minimum oxygen saturation (SPO2). After using Shapley analysis on our Logistic Regression model, it highlighted new top features that were not discussed in previous literature, such as bilirubin and maximum blood urea nitrogen. This information could be helpful to healthcare workers to see which patient features they need to monitor and evaluate.

While logistic regression exhibited the best performance, LightGBM, SVM, Random Forest, and XGBoost showed similar performance levels based on their AUC value and confidence intervals, as seen in Figure 6. This could mean that other models, when using accurate features and hyperparameters, are also able to accurately predict AKI in septic patients.

**Figure 6.**
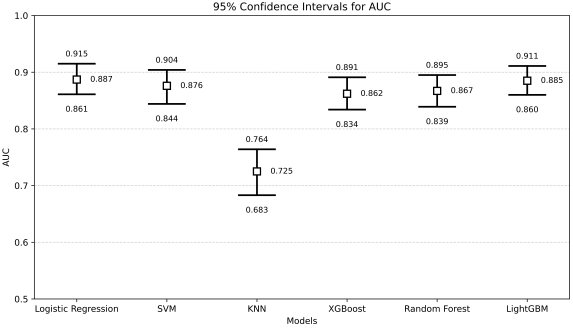
AUC with Confidence Intervals for each Model.

While the model shows a satisfactory result, some limitations cannot be overlooked. The data in the MIMIC-III database comes from the Beth Israel Deaconess Medical Center, a tertiary academic medical center located in Boston, Massachusetts. Using medical data from one single medical center might lead to some biased predictions in some particular areas like ethnicity, which were excluded during the process of our feature selection. Meanwhile, we cannot lose sight of the importance of ethnicity in the field of biological and medical research. Hence, some further improvements can be made by eliminating the bias caused by using a singular dataset. One study concluded that the diverse populations, results, and features are hindering the implementation of predictive models to aid septic patients with AKI [38]. The heterogeneity of medical patient data available will need to be addressed to improve the model’s performance and reliability going forward.

To enhance the clinical utility and robustness of AKI prediction, future research should test the model across diverse healthcare settings, expanding beyond the MIMIC-III dataset to improve generalizability. Future work could involve testing our model on diverse medical datasets to see if the results hold, providing a more inclusive model that considers potential demographic disparities. Additionally, integrating real-time monitoring could allow for continuous risk assessment, adapting to dynamic changes in patient conditions during ICU stays. Lastly, exploring advanced modeling techniques, such as ensemble learning or deep learning architectures, may offer further predictive power while maintaining interpretability for practical application. As more studies between sepsis and AKI are conducted, researchers could also test out new features to add or remove from the model to see if its performance increases. Furthermore, fine tuning the hyperparameters of our current model could also lead to improved AUC scores.

## Data Availability

All data produced in the present study are available upon reasonable request to the authors

## Acknowledgements

We acknowledge the use of the MIMIC-III database, maintained by the Laboratory for Computational Physiology at the Massachusetts Institute of Technology, which was instrumental in this study.

## Author contributions statement

Aleyeh Roknaldin conceptualized and implemented the machine learning models, conducted experiments, and performed model evaluations. Junyi Fan was responsible for drafting and refining the manuscript, ensuring clarity and coherence in the presentation of results. Shuheng Chen contributed to the generalizability analysis of the model, assessing its applicability across different patient cohorts. Elham Pishgar, Greg Placencia, and Maryam Pishgar provided critical insights into the study’s conceptual framework, ensuring the appropriateness of selected features, evaluation metrics, and overall study design. Maryam Pishgar supervised the research and provided guidance throughout the project. All authors reviewed and approved the final manuscript.

## Additional information

### Data Availability

The MIMIC-III dataset used in this study is publicly available at https://physionet.org/content/mimiciii/1.4/.

